# SRNT Health Equity Network Survey on Authentic Health Disparity/Equity Research

**DOI:** 10.1101/2024.03.04.24303279

**Authors:** Merideth Addicott, Josephine Hinds, Vita Mithi, George Kypriotakis, Wura Jacobs, Sydney A. Martinez, Rachel Denlinger-Apte, Lilianna Phan, Douglas E. Levy, Olatokunbo Osibogun, Kavita Mosalpuria, Danusha Selva Kumar, Lauren Czaplicki, Andy Tan

## Abstract

The Society for Research on Nicotine and Tobacco (SRNT) Health Equity Network (HEN) Evaluation Subcommittee members conducted an open-ended survey regarding what should be considered “authentic” health disparity/equity (HD/E) research and how the SRNT community defines this term. Anonymous surveys were emailed to over 300 SRNT HEN members, and invitees were asked to complete the survey if they conducted HD/E research or engaged in HD/E research in some other way. A total of 26 usable survey responses were collected and qualitatively coded. Respondents were asked to describe “authentic HD/E research”, challenges in their field, and indicators of good and poor quality HD/E research. Respondents expressed that authentic HD/E research investigates disparities/inequalities in health outcomes or access to healthcare services that are specific to communities defined by a social or demographic characteristic. Challenges included lack of funding, a slow rate of recruiting minority populations, and an under-valuation of HD/E research among funders and scientific journals. Indicators of good quality HD/E research were community involvement and a social justice context. Respondents also expressed concerns that poor quality HD/E research could inadvertently harm minoritized communities. As this field grows, we feel it is necessary for experts to set standards for the appropriate conduct of HD/E research, set benchmarks for success, and voice their concerns about the potentially negative impacts of poorly conducted HD/E research.

## Introduction

At the 2022 Society for Research on Nicotine and Tobacco (SRNT) annual conference, the Health Equity Theme Lecture was delivered by Linda A. Alexander, EdD, who gave a presentation titled “Authentic Equity in Tobacco Disparities Research”. Dr. Alexander sparked interest among SRNT Health Equity Network (HEN) members regarding what should be considered “authentic” health disparity/equity (HD/E) research and how the SRNT community defines this term. In her talk, Dr. Alexander joined other scholars in expressing concern that poorly conducted HD/E research could inadvertently advance negative stereotypes and further harm disadvantaged and minoritized populations. This raised the question of how researchers should judge quality in HD/E research.

## Methods

To address these questions and begin building consensus around these topics, the HEN Evaluation Subcommittee created a brief survey. Invitations to this anonymous survey were emailed to approximately 300 SRNT HEN members in September 2022. Invitees were asked to complete the survey if they conducted HD/E research or engaged in HD/E research in some other way. This project received an exemption and waiver of HIPAA authorization from the Wake Forest University School of Medicine Institutional Review Board (IRB00087203).

The survey consisted of five open-ended questions regarding HD/E research (enumerated below). A total of 26 completed survey responses were collected and qualitatively coded. Eighty percent of respondents reported conducting HD/E research, while the remainder reported engaging in some other way (e.g., collaborating with a primary HD/E researcher). Two authors (TH and VM) developed an a priori codebook based on the survey prompts and an initial review of responses. Then initial, deductive coding was conducted by sorting similar respondent answers within each survey question, based around questions asked that focused on “authenticity,” “priorities,” “challenges,” “quality indicators” and “clinical practice or health policy impacts,” respectively. From there, inductive subcodes were assigned to the categories that emerged from the first-level grouping for each question. Two coders each read the survey data and conducted the first-level coding independently. Then, coders discussed subcodes and grouping to reach a consensus. A third “tie breaker” coder was planned in the event of disagreement, though this was not necessary. See Table 1 for codes and subcodes. The themes that emerged from responses to the five survey questions are presented below.

**Table 1.** First-level, deductive coding was conducted based on anticipated answers to the open-ended questions. These a priori categories had themes that corresponded to direct prompts, such as how participants defined “authentic” and listed “priorities.” Within each category, subthemes emerged through deductive analysis and findings inside the groups of original primary themes. These were grouped in second and third-level sections, as demonstrated below.

|  |  |  |  |  |  |
| --- | --- | --- | --- | --- | --- |
| Survey Question : | *“How would you describe ‘authentic HD/HE research’ to someone in a different field of research?”* | *“How would you describe priority areas for HE/HE research concerning nicotine/tobacco use?”* | *“What kind of challenges exist in your field when conducting HD/HE research?”* | *“How would you describe indicators of good and poor quality in HD/HE research?”* | *“How do you think HD/HE research can impact clinical practice or health policy?”* |
| **1<sup>st</sup> Level Codes (General Themes)** | **<u>Authenticity</u>** | **<u>Priorities</u>** | **<u>Challenges</u>** | **<u>Quality Indicators</u>** | **<u>Impacts</u>** |
| 2<sup>nd</sup> Level Codes (within 1<sup>st</sup> level code) | ▪ According to purpose | ▪ Populations | ▪ “Upstream” Challenges | ▪ Good quality | ▪ General |
| 3<sup>rd</sup> Level / Granular Code (within 2<sup>nd</sup> level code) | ○ Seeking to understand/explain<br>○ seeking to address/fix<br>○ a standalone focus on a disparity population<br>○ seeking to dismantle systems | ○ Rural<br>○ Low SES<br>○ SGM communities<br>○ HIV+ pops<br>○ families with lower educational attainment<br>○ people with mental health issues | ○ funding<br>○ creating the “so what”<br>○ racism & bias<br>○ data gaps in existing research (scarcity, small samples) | ○ relationships with communities<br>○ research co-leads in the community<br>○ rigorous methods: clearly articulated research questions, rational<br>○ multi/trans-disciplinary<br>○ multilevel social justice considerations<br>○ actionable and feasible outcomes<br>○ community dissemination<br>○ rigorous intervention testing | ○ resource allocation and tailored initiatives<br>○ interdisciplinary: effects in public health, epidemiology, and researchers all communicating and integrating with front-line clinicians<br>○ |

| 2<sup>nd</sup> Level Codes (within 1<sup>st</sup> level code) | ▪ According to method | ▪ Behaviors | ▪ “Doing” Challenges | ▪ Poor quality | ▪ Clinical |
| --- | --- | --- | --- | --- | --- |
| 3<sup>rd</sup> Level / Granular Code (within 2<sup>nd</sup> level code) | ○ Involving community-based, culturally relevant approaches<br>○ led by member(s) of disparity populations<br>○ rigorous academic practices<br>○ uses an equity lens | ○ co-use of tobacco with cannabis, cocaine<br>○ co-use with harmful alcohol use<br>○ harm reduction | ○ competing priorities<br>○ authenticity and quality of researchers; rigor<br>○ connection & investment with communities<br>○ ensuring mutual benefit<br>○ recruitment | ○ low/no rigor<br>○ low/no community involvement<br>○ tokenism<br>○ descriptive findings without social justice implications | ○ Precision medicine |
| 2<sup>nd</sup> Level Codes (within 1<sup>st</sup> level code) |  | ▪ Outcomes | ▪ Application Challenges |  | ▪ Health Policy |
| 3<sup>rd</sup> Level / Granular Code (within 2<sup>nd</sup> level code) |  | ○ interventions<br>○ cultural tailoring<br>○ health communications<br>○ etiology: drivers, determinants, disparities<br>○ structural changes: policy/advocacy, creating political will<br>○ surveillance & measurement<br>○ capacity building: mentoring, training URM | ○ translating findings to action<br>○ harmful interpretations of research<br>○ intersectional inequities – how to address |  | ○ Policy-focused research: (un)intended consequences<br>○ Robust clinical trials |

## Results

### 1) How would you describe ‘authentic health disparity/equity research’ to someone in a different field of research?

Conceptually, respondents indicated that authentic HD/E research should seek to understand and/or explain disparities/inequities in health outcomes or access to healthcare services that are specific to communities defined by a sociodemographic characteristic, particularly, historically marginalized, minority communities. Respondents described how the goal of authentic HD/E research was to improve health outcomes or healthcare opportunities specific to the studied communities and to broadly dismantle systems that give rise to health disparities/inequities. Methodologically, respondents agreed on the need for community engagement in the research process, rather than “conducting research *on* them”, and approaches that consider and incorporate that community’s values or goals. Respondents also emphasized the need for rigorous academic practices and theories, as viewed through a lens of equity, to guide and conduct research. There were also concerns that the studied community should have an active voice and agency in the research so they can participate safely and the research is responsive to their needs.

Many respondents also directly addressed the use of the term “authentic”. While most considered this an important aspect of HD/E research, there were alternative viewpoints that suggested the term “authentic” may be an unsuitable or unscientific criterion. Some suggested that “authentic” could mean “well-informed” of the experience and perspectives of the studied community, and others indicated that “authentic” implied that the HD/E research was developed or led by member(s) of the studied community.

### 2) How would you describe priority areas for health disparity/equity research concerning nicotine/tobacco use?

Survey respondents identified a variety of populations and behaviors as priority areas for HD/E research concerning commercial nicotine/tobacco use. Identified populations included rural communities, those with low socioeconomic status and education levels, sexual and gender minorities, individuals living with HIV, and individuals with mental health problems. One respondent noted that all tobacco research is a priority since nearly all tobacco use now occurs among groups defined by socioeconomic status, race, identity, or co-morbid health concerns. Identified behaviors included poly-use of tobacco with other substances such as cannabis, cocaine, and alcohol, and the potential utility of harm reduction strategies.

Other identified priority areas included developing and implementing interventions tailored to specific communities that address disparities/inequities in a “holistic” fashion and improving surveillance and measurement of studied communities in national surveys. Several respondents also commented on the need to investigate the etiology of health disparities including the drivers and determinants of tobacco use, although one respondent expressed concern regarding the “use and misuse of genetic studies to examine racial/ethnic differences”. Some respondents noted prioritizing “political will” to enact policies (e.g., reduce targeted tobacco industry advertising; ban flavors), and a broader need for more mentoring and training of researchers from under-represented/marginalized backgrounds.

### 3) What kind of challenges exist in your field when conducting health disparity/equity research?

Respondents identified several upstream challenges to conducting HD/E research, like funding challenges and difficulties in addressing the “so what?” factor in order to be funded and published.

Several respondents lamented the structural racism and bias in academia and medicine, suggesting that HD/E research is undervalued and under-recognized. Others noted the gaps in existing datasets regarding racial minorities, and many articulated the difficulties of recruiting racial minorities (e.g., slow rate of recruitment, challenges engaging with the population) that often result in small sample sizes, which impacts data analysis.

Methodological challenges included a lack of diversity and training among researchers and study staff in conducting HD/E research, as well as competing priorities among grant funders and scientific journals. There were several comments on the complexities of connecting with the studied community and the lack of “time to devote to recruitment”. Respondents specifically highlighted how it takes time to build trust and to ensure mutual benefit between the researchers and the studied community.

Respondents also indicated challenges in the application of HD/E research, including issues in the translation of outcomes in research into actionable reductions in disparities, and difficulties in addressing inequities when there are multiple contributing factors. Several respondents were concerned that some investigators may simply include results broken down along racial/ethnic lines without appreciating the impact on the population, which could perpetuate harmful stereotypes among already minoritized communities.

### 4) How would you describe indicators of good and poor quality in health disparity/equity research?

Respondents agreed that an important indicator of good quality HD/E research was evidence of strong relationships between the researchers and the studied communities, with members of the community co-leading and/or “power-sharing” the research. Other indicators of good quality were rigorous methods with clearly articulated research questions and theory-driven rationale and use of multi/trans-disciplinary methods. Respondents also advocated for incorporating multilevel social justice context within good HD/E research, in addition to actionable and feasible research outcomes that empower communities. Respondents expressed that rigorous methods should be used to test interventions and results should always be disseminated among the studied community.

Some markers of poor quality were little to no community involvement or community input into the project. Several respondents warned against research that provides descriptive findings of differences among minority subpopulations without any social justice implications, likening it to a discussion of “race differences without discussing the depth of systemic racism.”

### 5) How do you think health disparity/equity research can impact clinical practice and/or health policy?

Broadly, respondents felt that HD/E research could, meaningfully impact clinical and health policy, and it is up to researchers to “highlight the importance of understanding and addressing” inequities for population and individual health outcomes. Respondents described how HD/E research could influence policy by encouraging resource allocation and tailored initiatives for underserved communities. Other respondents believed that as an interdisciplinary science, HD/E research on public health and epidemiology needs to be communicated with front-line clinicians to improve standard of care. One respondent emphasized that HD/E research should seek to put pressure on politicians to support universal healthcare as a social justice goal. Some respondents suggested there were potential clinical impacts in helping determine what intervention is most effective for different groups since, historically, biomedical research focused on White men. Several also noted health policy impacts such as the importance of policy-focused research to uncover potentially negative unintended consequences of current clinical practice, and the need for robust, well-powered clinical trials to inform good quality policy.

## Discussion

Experts conducting HD/E research were asked to describe “authentic health disparity/equity research”, challenges in their field, and indicators of good and poor quality HD/E research. Responses revealed several cross-cutting themes. Respondents felt strongly that HD/E research needs community-based approaches that must be informed by social justice. Rigor in HD/E research is important, and HD/E research outcomes should be actionable and ultimately help reduce disparities. Based on the themes discussed, we propose an initial definition of authentic HD/E research: the investigation of disparities/inequities in health outcomes or access to healthcare services that are specific to communities defined by a social or demographic characteristic, in particular, historically marginalized, minority communities. Importantly, authentic HD/E research actively involves the studied communities in the research process, with the specific goal of improving the health of the studied community and the broader goal of dismantling systems that give rise to health disparities/inequities.

Some limitations of this survey include the small sample size and the lack of information regarding respondents’ level of expertise. Furthermore, there was a lack of specificity in the survey questions and there was no opportunity to delve more deeply into ambiguous responses.

Our approach and commentary are motivated by a history in which minoritized communities have been targeted by the tobacco industry and have experienced more relaxed tobacco control regulations. Combined with less healthcare access, tobacco-related health disparities must be identified and mitigated for the sake of social justice and to improve health for all populations (Sheffer et al., 2022, Tan et al., 2023). As this field grows, experts should continue to inform practices for the appropriate conduct of HD/E research, set benchmarks for success, and voice their concerns about the potentially negative impacts of poorly conducted HD/E research. While there was a lack of specificity in the questions and no opportunity to delve more deeply into ambiguous responses, we hope the results of this initial work inspire future discussions on these topics and hypothesis-generating research, help the larger community build consensus on these themes, and ultimately advance health equity in tobacco control.

## Data Availability

Due to the sensitive nature of the opinions in the survey, and the potential for survey respondents to be identified based on their answers, this survey data is not available for sharing.

## Acknowledgments

We would like to thank Dr. Linda Alexander for giving the presentation on *“Authentic Equity in Tobacco Disparities Research”* at the 2022 SRNT Annual Conference meeting, the members of the SRNT community, and the conference organizers.

## Notes

Declaration of Interests: The authors have no conflicts to declare.

### Competing Interest Statement

The authors have declared no competing interest.

### Funding Statement

This study did not receive any funding.

### Author Declarations

This project received an exemption and waiver of HIPAA authorization from the Wake Forest University School of Medicine Institutional Review Board (IRB00087203).

